# Obesity, hypertension and tobacco use associated with left ventricular remodelling and hypertrophy in South African Women: Birth to Twenty Plus Cohort

**DOI:** 10.1101/2022.03.14.22272334

**Authors:** Andrea Kolkenbeck-Ruh, Larske M. Soepnel, Simone H. Crouch, Sanushka Naidoo, Wayne Smith, Shane A. Norris, Justine Davies, Lisa J. Ware

**Affiliations:** SAMRC/Wits Developmental Pathways for Health Research Unit, Faculty of Health Sciences, University of the Witwatersrand, Johannesburg 2000, South Africa; Julius Global Health, Julius Centre for Health Sciences and Primary Care, University Medical Centre. Utrecht, Utrecht University, Utrecht, The Netherlands; Hypertension in Africa Research Team (HART), North-West University, South Africa; South African Medical Research Council: Unit for Hypertension and Cardiovascular Disease, North-West University, South Africa; School of Health and Human Development, University of Southampton, Southampton, UK; Institute of Applied Health Research, University of Birmingham, Birmingham, UK; DSI-NRF Centre of Excellence in Human Development, University of the Witwatersrand, Johannesburg 2000, South Africa

**Keywords:** left ventricular hypertrophy, ventricular remodelling, Sub-Saharan Africa, Child health, Women’s health

## Abstract

**Background:** Left ventricular hypertrophy (LVH) is a known marker of increased risk in developing future life-threating CVD, though it is unclear how health risk factors, such as obesity, blood pressure and tobacco use, associate with left ventricular (LV) remodelling and LVH across generations of urban African populations.

**Methods:** Black female adults (n=123; age: 29-68 years) and their children (n=64; age: 4-10; 55% female) were recruited from the Birth to Twenty Plus Cohort in Soweto, South Africa. Tobacco and alcohol use, physical activity, presence of diabetes mellitus, heart disease and medication were self-reported. Height, weight, and blood pressure were measured in triplicate. Echocardiography was used to assess LV mass at end-diastole, perpendicular to the long axis of the LV and indexed to body surface area to determine LVH.

**Results:** Hypertension and obesity prevalence were 35.8% and 59.3% for adults and 45.3% and 6.3% for children. Self-reported tobacco use in adults was 22.8%. LVH prevalence was 35.8% (n=44) in adults (75% eccentric; 25% concentric), and 6.3% (n=4) in children (all eccentric). Prevalence of concentric remodelling was 15.4% (n=19) in adults and observed in one child. In adults, obesity (OR: 2.54 (1.07-6.02; p=0.02) and hypertension (3.39 (1.08-10.62; p=0.04) significantly increased the odds of LVH, specifically eccentric LVH, while concentric LVH was associated with self-reported tobacco use (OR: 4.58 (1.18-17.73; p=0.03; n=11). Although no logistic regression was run within children, of the four children LVH, three had elevated blood pressure and the child with normal blood pressure was overweight.

**Conclusions:** The association between obesity, hypertension, tobacco use and LVH in adults, and the 6% prevalence of LVH in children, calls for stronger public health efforts to control risk factors and monitor children at who are risk.

## Background

Despite medical advances, cardiovascular disease (CVD) remains a global health concern affecting more than 37 million individuals [1]. CVD are a leading cause of death globally [2–4]. CVD risk factors, such as hypertension, obesity, type II diabetes mellitus (T2DM), smoking, dyslipidaemia, a sedentary lifestyle, unhealthy diets and excess alcohol intake [4, 5], are increasing in sub-Saharan Africa. South Africa is no exception with rising prevalence of known CVD risk factors, largely as a consequence of urbanisation and marked changes in health behaviour [6–8]. Thus prevention of CVD is an important national public health goal.

Heart failure is the dominant form of CVD in South Africa, of which the main cause is hypertension [9]. Unlike high-income countries, in South Africa heart failure typically affects younger working individuals, and has an in-hospital mortality rate of approximately 8.4%, while 6 months post hospital discharge, fatality rate stands at approximately 26% [10]. As a result of the impact on young, economically active individuals, as well as the high fatality rate, heart failure in South Africa has a disproportional impact on an already fragile economy.

A key aspect of the pathological process of CVD, ultimately leading to heart failure and stroke [11, 12], is cardiac remodelling. The term is most commonly applied to the left ventricle (LV) in which the chamber changes in size, shape or structure [13]. Left ventricular hypertrophy (LVH), defined as an increase in muscle mass of the LV, is associated with a four-fold increase in heart failure [14–20]. As a result, regression of LVH is a goal of cardiovascular risk reduction [21–24]. Many of the risk factors contributing to the systemic and regional hemodynamic changes that drive the development of LVH [25], are modifiable, including obesity [26], tobacco and alcohol use [27] and elevated blood pressure and hypertension [26].

In South African adult women, there has been a sharp rise in prevalence of overweight/obesity (68%), hypertension (46%), regular alcohol use (17%) [28] and current tobacco smokers (7.6%) [29]. Given the increasing CVD rates, and the restricted access to echocardiography in the public health sector [30], understanding the role of health risk factors in LVH development may have major implications for preventing LVH or interventions for regression of LVH. Furthermore, the growing burden of cardiovascular risk factors could lead to growing burden of LVH and potential increase the burden in offspring. Therefore, the aim of the study was to identify the prevalence of LVH as well as the health factors associated with LV remodelling and LVH, within black South African adult women and their pre-pubescent children to inform public health efforts to promote cardiovascular health.

## Methods

### Study population

Data was collected during a cross-sectional study of vascular health in an existing birth cohort known as the Birth to Twenty Plus (BT20+) Cohort, which started in 1990 (described previously [31]) and now includes three generations. The aim of this study was to investigate the intergenerational transmission of cardiovascular health; the BT20 index female (known as 2^nd^ generation or 2G; aged 28-29 years); their birth children or 3^rd^ generation (3G, aged 4-10 years, female or male); and their biological mothers or 1^st^ generation (1G, aged 43-68 years, female). For the purpose of the present study, the first and second-generation participants were combined and classified as the “adult group”. Thus, 79 families, 237 participants (n=158 adults and n=79 children) were invited to attend for cardiovascular assessment where left ventricular remodelling and hypertrophy were assessed using echocardiography. All data were collected at the Developmental Pathways for Health Research Unit, at Chris Hani Baragwanath Academic Hospital in Soweto. Of the 237 participants recruited, n=187 (78.9%) were included in analysis (**Figure 1**). Trained researchers who spoke the participant’s home language explained the study and all participants gave written informed consent (adults) or assent (children aged 7-10 years) prior to data collection. The Human Research Ethics Committee (Medical) of the University of the Witwatersrand approved the protocol (M190263).

**Figure 1.**
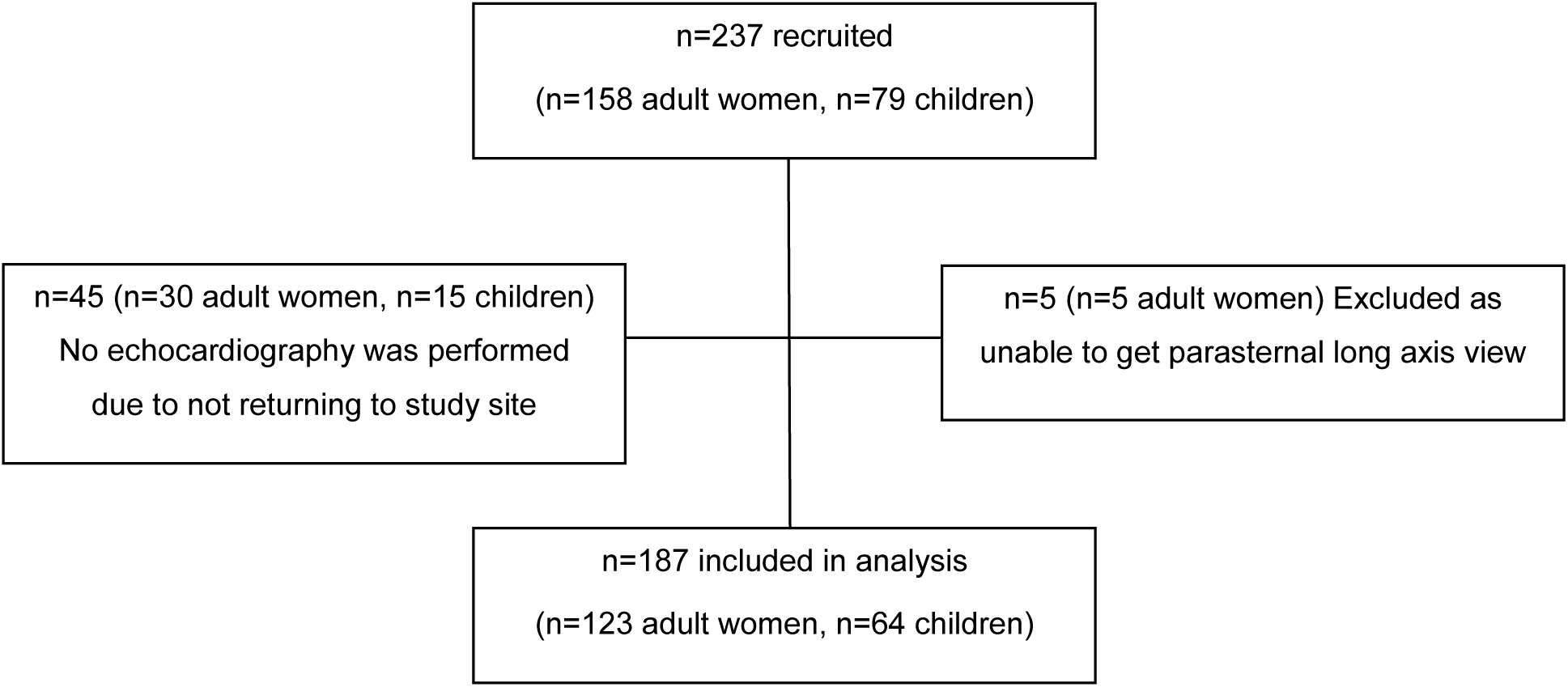
Study flow diagram

### Health risk factors

Tobacco use was assessed using the Global Adult Tobacco Survey [32], alcohol use with the WHO Alcohol Use Disorders (WHO-AUDIT) test [33] and physical activity using the Global Physical Activity Questionnaire (GPAQ) [34]. Self-reported medical history (diabetes mellitus [T2DM), hypertension, high cholesterol, or previous heart disease] and current medication use) was assessed by questionnaire. Thereafter, anthropometry, including height, weight and waist circumference, was measured in triplicate using standardised World Health Organization measurement protocols [35]. Brachial blood pressure (BP) was measured using automated devices (MIT5 for adults; HBP-1300 for children [36]; Omron Healthcare, Kyto, Japan). Following the International Society of Hypertension (ISH) measurement guidelines [37], participants were seated and asked to rest for at least 5-minutes prior to measurement of BP in their right arm three times in a quiet room with the monitor facing away from the participant; there was a 2-minute rest interval between the measurements. The first BP measurement was discarded and the second and third measurements were averaged for analysis.

### Echocardiography

Transthoracic echocardiography examination was performed by a single echocardiographer, with the Mindray DC-70 Ultrasound system (Mindray, Shenzen China). Measurements of chamber dimensions were taken from 2D mode and left ventricular (LV) mass and relative wall thickness (RWT) were calculated. Linear measurements were made according the American Society of Echocardiography (ASC) Guidelines: Recommendations for Cardiac Chamber Quantification in Adults [38]. LV mass was assessed at end-diastole perpendicular to the long axis of the LV. LVM was calculated according to the Devereus formula: *LVM (g) = 0*.*8 × 1*.*04 ((LVDd+IVSd+LVPWd)*^*3*^ *– LVDd*^*3*^*) + 0*.*6* where LVDd = left ventricular diastolic diameter; IVSd = intraventricular septal diameter, LVPWd = left ventricular posterior wall thickness in diastole [38]. The left ventricular mass index (LVMI) was calculated as a ratio of LVM indexed to body surface area [38]. The RWT was defined as: *2 x LVPWd/LVDd*, where LVPWd = left ventricular posterior wall thickness in diastole; LVDd = left ventricular diastolic diameter [38]. RWT was used to categorise LV mass and the pattern of remodelling. Left ventricular hypertrophy (LVH) was defined as LVMI >95 g/m^2^ for adult women [38] and LVMI >95^th^ percentile (88.9 g/m^2^) for children [39], and eccentric LVH (LVH with RWT ≤0.42) and concentric LVH (LVH with RWT >0.42) as well as concentric remodelling (normal LVMI with RWT >0.42) [38].

### Data Management

In adults, body mass index (BMI; weight (kg)/height (m)^2^) was categorised according to the World Health Organisation (WHO) classification as follows: <18.5 kg/m^2^ as underweight; 18.5-24.9 kg/m^2^ as normal; 25-29.9 kg/m^2^ as overweight; and ≥30 kg/m^2^ as obese [40]. In children (>5 years), age adjusted z-scores for BMI were calculated using the WHO reference and children were categorised as overweight if their BMI z-score was between 1-2, and obese if their BMI z-score was >2 [40]. While in children aged four years, age adjusted z-scores for BMI were categorised as overweight if their BMI z-score was between 2-3, and obese if their BMI z-score was >3 [41]. In adults, hypertension was defined as a BP ≥140 mmHg systolic or ≥90 mmHg diastolic or currently taking anti-hypertensive medication; in those who were not on anti-hypertensive medication elevated BP was defined as a BP 130-139 mmHg systolic or 85-89 mmHg diastolic, following the International Society of Hypertension (ISH) Global Hypertension Practice Guidelines [37]. For children, elevated BP was defined as BP ≥90^th^ percentile for height and age, following the American Academy of Paediatrics Clinical Practice 2017 guidelines [42].

### Statistical analysis

SPSS statistics 25.0 (IBM, Chicago, IL, USA) was used for statistical analysis. Non-normality of data was confirmed using visual inspection of histograms and the Shapiro–Wilk test. Multivariable logistic regression, after checking for multicollinearity by confirming a variance inflation factor of less than five [43], was conducted to examine the relationship between LVH / LV remodelling and the following health factors: age, moderate-vigorous physical activity, sedentary behaviour, obesity, hypertension, previous diagnosis of T2DM, high cholesterol, ever tobacco use and alcohol use. Statistical significance for all analyses was set at p<0.05. Due to the low prevalence of LVH in children, regression analysis was only performed in adults.

## Results

### Characteristics of the participants

The median age for adult women was 41 years (range 28 to 68 years, n=123) while for children it was 7 years (range 4 to 10 years, n=64, 54.7% female) **(Table 1)**. Median BMI was 32.5 kg/m^2^ (IQR: 26.6-37.7 kg/m^2^) and 15.8 kg/m^2^ (IQR: 14.8-17.2 kg/m^2^) for adults and children respectively, with 59.3% of adults and 6.3% of children classified as obese. Elevated blood pressure (BP) on measurement was found in 41.5% of adults. While 35.8% of adults were classified as hypertensive with 75% on treatment, primarily a diuretic. Nearly sixty percent (59.4%) of children had elevated BP. Of the 123 adults, 22.8% were current or past tobacco users (smoking and smokeless tobacco) and 55.3% regularly consumed alcohol. Three adults reported having previous heart disease. A small percentage of adults reported having high cholesterol and T2DM (12.2% and 2.4% respectively). While all participants with self-reported diabetes were on treatment, only 26.7% of adults with high cholesterol were.

**Table 1.**
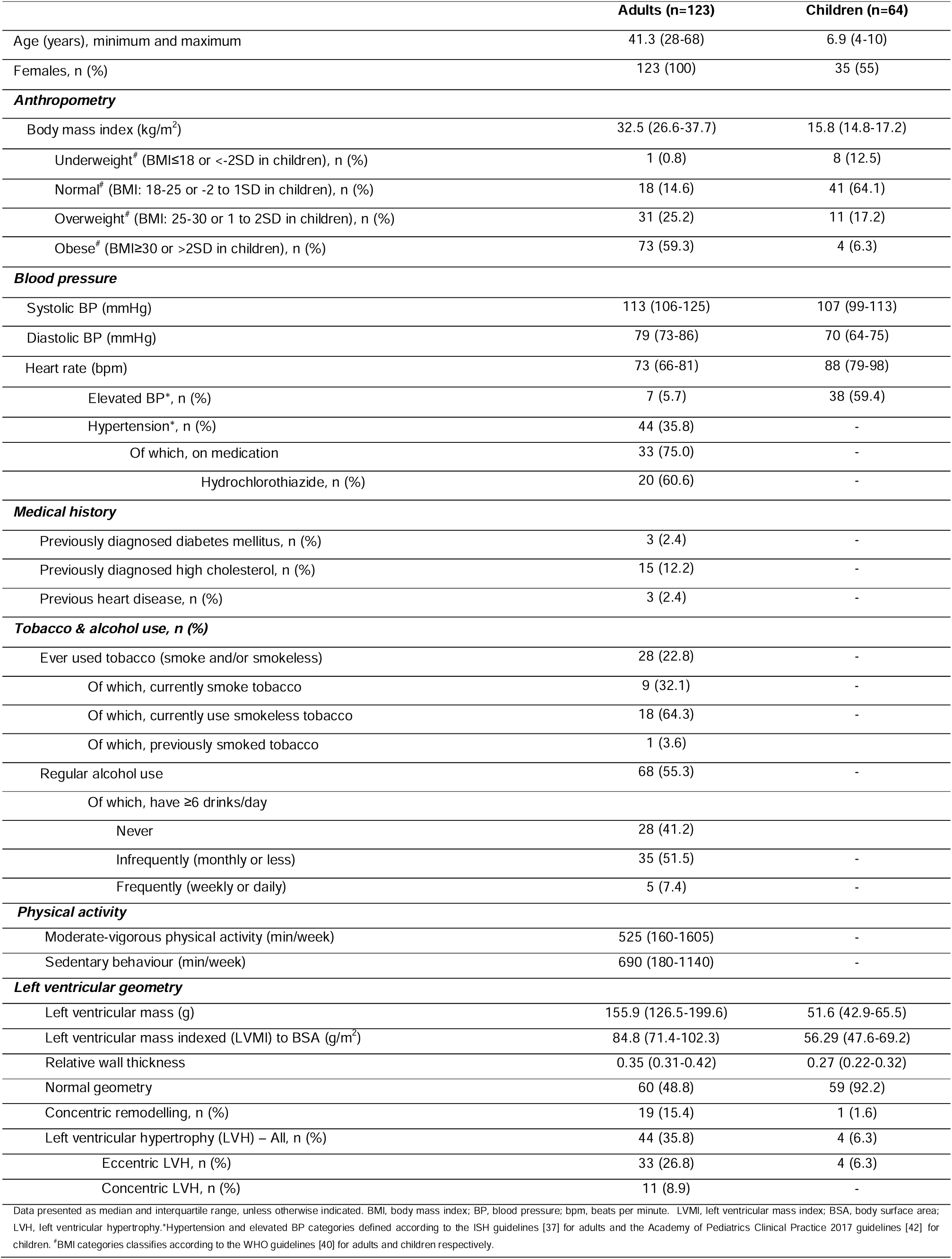
Characteristics of the adults and children (n=187)

|  | Adults (n=123) | Children (n=64) |
| --- | --- | --- |
| Age (years), minimum and maximum | 41.3 (28-68) | 6.9 (4-10) |
| Females, n (%) | 123 (100) | 35 (55) |
| **Anthropometry** |  |  |
| Body mass index (kg/m<sup>2</sup>) | 32.5 (26.6-37.7) | 15.8 (14.8-17.2) |
| Underweight<sup>#</sup> (BMI≤18 or <-2SD in children), n (%) | 1 (0.8) | 8 (12.5) |
| Normal<sup>#</sup> (BMI: 18-25 or -2 to 1SD in children), n (%) | 18 (14.6) | 41 (64.1) |
| Overweight<sup>#</sup> (BMI: 25-30 or 1 to 2SD in children), n (%) | 31 (25.2) | 11 (17.2) |
| Obese<sup>#</sup> (BMI≥30 or >2SD in children), n (%) | 73 (59.3) | 4 (6.3) |
| **Blood pressure** |  |  |
| Systolic BP (mmHg) | 113 (106-125) | 107 (99-113) |
| Diastolic BP (mmHg) | 79 (73-86) | 70 (64-75) |
| Heart rate (bpm) | 73 (66-81) | 88 (79-98) |
| Elevated BP*, n (%) | 7 (5.7) | 38 (59.4) |
| Hypertension*, n (%) | 44 (35.8) | - |
| Of which, on medication | 33 (75.0) | - |
| Hydrochlorothiazide, n (%) | 20 (60.6) | - |
| **Medical history** |  |  |
| Previously diagnosed diabetes mellitus, n (%) | 3 (2.4) | - |
| Previously diagnosed high cholesterol, n (%) | 15 (12.2) | - |
| Previous heart disease, n (%) | 3 (2.4) | - |
| **Tobacco & alcohol use, n (%)** |  |  |
| Ever used tobacco (smoke and/or smokeless) | 28 (22.8) | - |
| Of which, currently smoke tobacco | 9 (32.1) | - |
| Of which, currently use smokeless tobacco | 18 (64.3) | - |
| Of which, previously smoked tobacco | 1 (3.6) | - |
| Regular alcohol use | 68 (55.3) | - |
| Of which, have ≥6 drinks/day |  |  |
| Never | 28 (41.2) |  |
| Infrequently (monthly or less) | 35 (51.5) | - |
| Frequently (weekly or daily) | 5 (7.4) | - |
| **Physical activity** |  |  |
| Moderate-vigorous physical activity (min/week) | 525 (160-1605) | - |
| Sedentary behaviour (min/week) | 690 (180-1140) | - |
| **Left ventricular geometry** |  |  |
| Left ventricular mass (g) | 155.9 (126.5-199.6) | 51.6 (42.9-65.5) |
| Left ventricular mass indexed (LVMI) to BSA (g/m<sup>2</sup>) | 84.8 (71.4-102.3) | 56.29 (47.6-69.2) |
| Relative wall thickness | 0.35 (0.31-0.42) | 0.27 (0.22-0.32) |
| Normal geometry | 60 (48.8) | 59 (92.2) |
| Concentric remodelling, n (%) | 19 (15.4) | 1 (1.6) |
| Left ventricular hypertrophy (LVH) – All, n (%) | 44 (35.8) | 4 (6.3) |
| Eccentric LVH, n (%) | 33 (26.8) | 4 (6.3) |
| Concentric LVH, n (%) | 11 (8.9) | - |

Data presented as median and interquartile range, unless otherwise indicated. BMI, body mass index; BP, blood pressure; bpm, beats per minute. LVMI, left ventricular mass index; BSA, body surface area; LVH, left ventricular hypertrophy.\*Hypertension and elevated BP categories defined according to the ISH guidelines [37] for adults and the Academy of Pediatrics Clinical Practice 2017 guidelines [42] for children. <sup>#</sup>BMI categories classifies according to the WHO guidelines [40] for adults and children respectively.

The median LVMI was 84.75 (IQR: 71.36-102.33) g/m^2^ and 56.29 (IQR: 47.61-69.17) g/m^2^ for adults and children, respectively **(Table 1)**. Within adults, 48.8% had normal LV geometry, while15.4% had concentric remodelling. The remaining 35.8% had LVH, of which 26.8% had eccentric LVH and 8.9% had concentric LVH. In contrast, 92.2% of children had normal LV geometry, 1.6% had concentric remodelling, 6.3% had eccentric remodelling only, and no concentric LVH was observed.

### Left ventricular (LV) remodelling and left ventricular hypertrophy (LVH) in adults

**Table 2**, compares characteristics of adults with and without LVH. Overall, women with LVH were significantly older (48 (min: 29, max 66) vs 29 (min: 28, max: 68) years, p=0.04), were more obese (73% vs 52%, p=0.04), and were hypertensive (52% vs 27%, p=0.01) compared to those without LVH. When considering LVH regardless of pattern, after adjusting for age, moderate-vigorous physical activity, sedentary behaviour, obesity, hypertension, T2DM, high cholesterol, ever tobacco use and regular alcohol use, hypertension (OR: 3.39 (1.08-10.62), p=0.04) and obesity (OR: 2.54 (1.07-6.02) were significantly associated with LVH. Obesity was the only factor significantly associated with eccentric LVH (OR: 3.46 (1.28-9.37), p=0.02), and reported ever tobacco use was the only factor significantly associated with concentric LVH (OR: 5.06 (1.24-20.69), p=0.02), although the number of adults with this condition was small. No other significant associations were found with LV remodelling in the multivariable logistic regression models (**Table 3**).

**Table 2.** Sociodemographic characteristics of the women adults with and without left ventricular hypertrophy

|  | With LVH | Without LVH | p-value |
| --- | --- | --- | --- |
| n | 44 | 79 | - |
| Age (years), minimum and maximum | 48 (29,66) | 29 (28,68) | **0.04** |
| **Anthropometry** |  |  |  |
| Body mass index (kg/m<sup>2</sup>) | 34.9 (28.9-40.4) | 30.3 (25.9-34.6) | **0.01** |
| Underweight<sup>#</sup> (BMI≤18), n (%) | 1 (2.3) | 0 | - |
| Normal<sup>#</sup> (BMI: 18-25) n (%) | 5 (11.4) | 13 (16.5) | 0.10 |
| Overweight<sup>#</sup> (BMI: 25-30), n (%) | 6 (13.6) | 25 (31.60) | **0.03** |
| Obese<sup>#</sup> (BMI≥30), n (%) | 32 (72.7) | 41 (51.9) | **0.04** |
| **Blood pressure** |  |  |  |
| Systolic BP (mmHg) | 119 (108-129) | 111 (104-123) | 0.16 |
| Diastolic BP (mmHg) | 74 (72-90) | 79 (73-84) | 0.99 |
| Heart rate (bpm) | 56 (65-81) | 74 (66-82) | 0.55 |
| Elevated BP*, n (%) | 3 (6.8) | 4 (5.1) | 0.70 |
| Hypertension*, n (%) | 23 (52.3) | 21 (26.6) | **0.01** |
| Of which, on medication | 16 (69.6) | 17 (80.9) | **0.001** |
| Hydrochlorothiazide, n (%) | 12 (75.0) | 8 (47.1) | **0.02** |
| **Medical history** |  |  |  |
| Previously diagnosed diabetes mellitus, n (%) | 3 (6.8) | 0 | 0.09 |
| Previously diagnosed high cholesterol, n (%) | 6 (13.6) | 9 (11.4) | 0.48 |
| Previous heart disease, n (%) | 1 (2.3) | 0 | - |
| **Tobacco & alcohol use, n (%)** |  |  |  |
| Ever used tobacco (smoke and/or smokeless) | 13 (29.5) | 15 (19.0) | 0.19 |
| Of which, currently smoke tobacco | 4 (9.1) | 5 (6.30) | 0.72 |
| Of which, currently use smokeless tobacco | 8 (18.2) | 10 (12.7) | 0.43 |
| Of which, previously smoked tobacco | 1 (2.5) | 0 | 0.35 |
| Ever used alcohol | 23 (52.3) | 45 (57.0) | 0.71 |
| Of which, have ≥6 drinks/day |  |  |  |
| Never | 11 (47.8) | 17 (37.8) | **0.04** |
| Infrequently (monthly or less) | 10 (43.5) | 25 (55.6) | **0.04** |
| Frequently (weekly or daily) | 2 (8.7) | 3 (6.7) | 0.06 |
| **Physical activity** |  |  |  |
| Moderate-vigorous physical activity (min/week) | 600 (60-1823) | 510 (180-1215) |  |
| Sedentary behaviour (min/week) | 509 (19-960) | 720 (210-1320) |  |

Data presented as median and interquartile range, unless otherwise indicated. BMI, body mass index; BP, blood pressure; bpm, beats per minute; LVH, left ventricular hypertrophy. \*Hypertension and elevated BP categories defined according to the ISH guidelines [37]. <sup>#</sup>BMI categories classifies according to the WHO guidelines [40]. p<0.05 was considered significant.

**Table 3.** Multivariable adjusted associations between health risk factors and left ventricular remodelling and left ventricular hypertrophy in adults

| Models with → | LV remodelling<br>n=19 |  | LVH (all patterns)<br>n=44 |  | Eccentric LVH<br>n=33 |  | Concentric LVH<br>n=11 |  |
| --- | --- | --- | --- | --- | --- | --- | --- | --- |
|  | OR (95%CI) | p-value | OR (95%CI) | p-value | OR (95%CI) | p-value | OR (95%CI) | p-value |
| Age | 0.99 (0.94-1.05) | 0.77 | 0.99 (0.95-1.04) | 0.76 | 0.98 (0.94-1.03) | 0.37 | 1.03 (0.96-1.10) | 0.46 |
| MVPA | 1.00 (0.99-1.00) | 0.33 | 1.00 (1.00-1.00) | 0.29 | 1.00 (1.00-1.00) | 0.37 | 1.00 (1.00-1.001) | 0.62 |
| Sedentary behaviour | 1.00 (1.00-1.001) | 0.42 | 1.00 (0.99-1.00) | 0.47 | 1.00 (0.99-1.00) | 0.25 | 1.00 (1.00-1.001) | 0.49 |
| Obesity<sup>*</sup> | 0.58 (0.19-1.73) | 0.33 | 2.54 (1.07-6.02) | **0.03\*** | 3.46 (1.28-9.37) | **0.02\*** | 1.03 (0.24-4.32) | 0.97 |
| Hypertension<sup>#</sup> | 3.44 (0.80-14.82) | 0.09 | 3.39 (1.08-10.62) | **0.04\*** | 3.83 (1.10-13.31) | 0.03 | 1.23 (0.17-8.96) | 0.84 |
| T2DM<sub>...</sub> | 1.92 (0.47-7.78) | 0.36 | 1.05 (0.29-3.86) | 0.94 | 0.82 (0.18-3.72) | 0.79 | 1.41 (0.22-9.21) | 0.72 |
| High cholesterol<sub>...</sub> | 4.56 (0.50-41.24) | 0.18 | 1.33 (0.39-4.55) | 0.65 | 1.61 (0.42-6.20) | 0.49 | 0.73 (1.00-5.40) | 0.76 |
| Tobacco<sup>§</sup> use<sub>...</sub> | 1.14 (0.34-3.84) | 0.83 | 1.87 (0.73-4.80) | 0.192 | 0.84 (0.29-2.41) | 0.75 | 5.06 (1.24-20.69) | **0.02\*** |
| Alcohol use<sub>...</sub> | 1.33 (0.44-3.99) | 0.61 | 1.22 (0.51-2.94) | 0.656 | 0.74 (0.29-1.92) | 0.54 | 3.38 (0.68-16.79) | 0.14 |

OR, odds ratio; CI, confidence interval; MVPA, moderate-vigorous physical activity; LV, left ventricular; LVH, left ventricular hypertrophy; T2DM, type 2 diabetes mellitus. <sup>\*</sup> As defined as a body mass index (BMI) $\geq 30$ kg/m<sup>2</sup> according to the WHO guidelines [40]. <sup>#</sup> Hypertension as defined by the ISH guidelines [37]. Self-reported. <sup>§</sup> Smoking and smokeless tobacco. \* p<0.05 was considered significant.

**Figure 2**, depicts those adults with LVH and the number of health factors. It was observed the prevalence of LVH in adults significantly increased (p=0.002) with the number of health risk factors reported.

**Figure 2.**
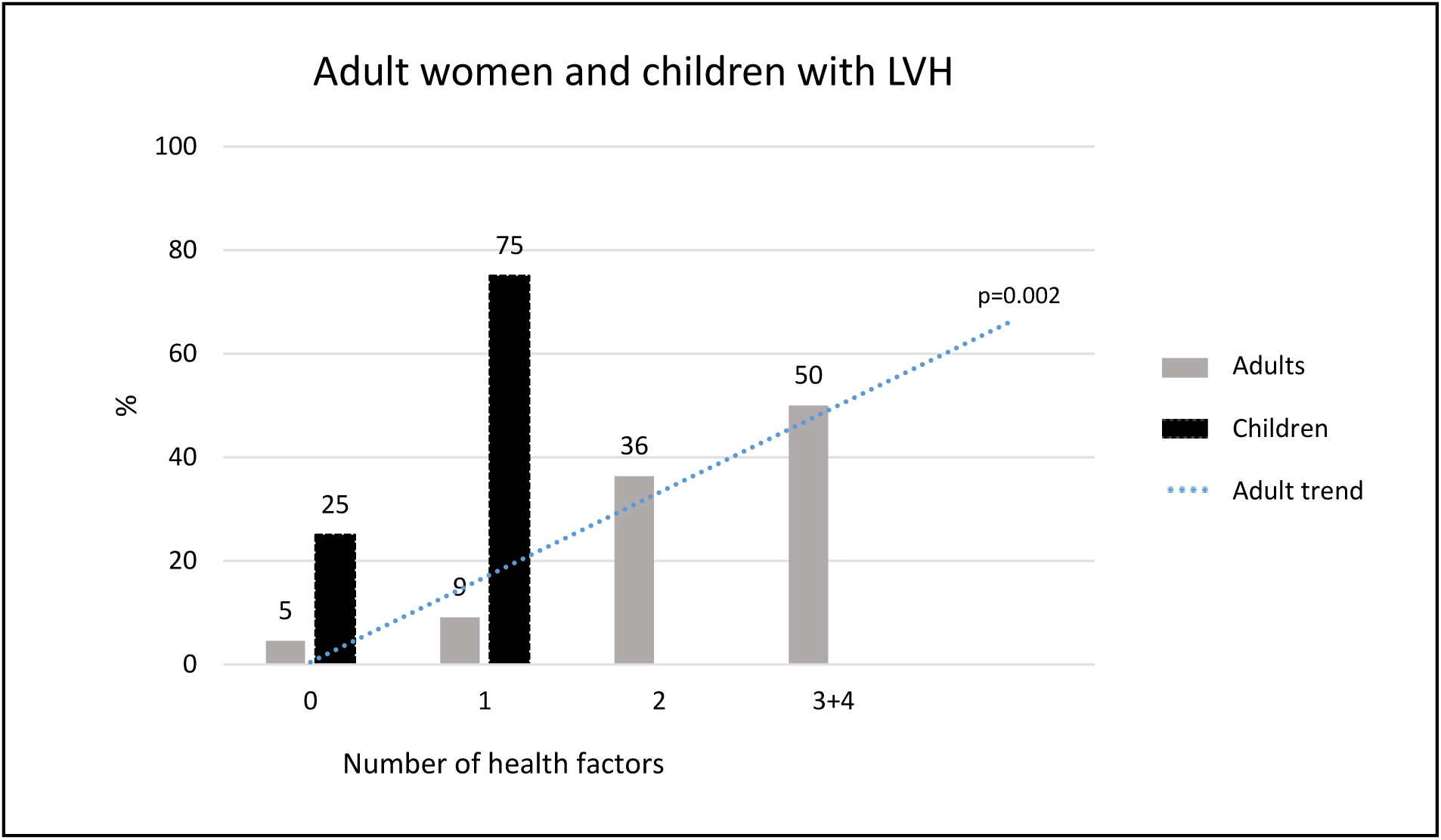
Percentage of South African adult women and children with left ventricular hypertrophy (LVH) according to the number of health factors. Health behaviours for women included: obesity, hypertension, self-reported type II diabetes mellitus, self-reported high cholesterol, tobacco use, and alcohol consumption. Health behaviours for children included: obesity, elevated blood pressure, whether their mother and or grandmother use tobacco.

### Left ventricular (LV) remodelling and left ventricular hypertrophy (LVH) in children

Over 90% of children (median age 7 years, range 4-10, 59.3% female) had normal LV geometry despite 59.4% (n=38) having elevated BP, 45.3.% (n=26) being hypertensive and 23.4% (n=15) being overweight or obese (**Table 1**). Of the four children with LVH (all eccentric LVH), three had elevated BP and were normal weight, while the fourth child had normal BP, however, was overweight. **Figure 2**, depicts those children with LVH and the number of health factors, in which 3 out of the 4 children with LVH has one health factor.

## Discussion

In the present study, we assessed the prevalence of LVH in black South African women and their children/grandchildren and health factors associated with LVH. We found a high prevalence of LVH in South African black adult women (28-68 years), with over a quarter having eccentric LVH. Obesity and hypertension, two modifiable health factors, were associated with eccentric LVH, while self-reported tobacco use was related to concentric LVH. Of the women with LVH, half of the women had three or four health risk factors. Despite the small sample size, 6% of children aged 4-10 years had eccentric LVH, of which three had elevated blood pressure (BP). The current study highlights that multiple factors significantly drive the development of LVH in adult women. In addition, although only four children had LVH, the high prevalence of elevated BP and children is concerning and aids in supporting ongoing calls for prevention efforts, particularly for youth.

In African countries, CVD is increasing rapidly. LVH is a known marker of increased risk for developing future life-threating CVD and there is evidence to suggest that LVH is higher in black populations of African descent [44]. Various studies conducted over the past decade in Sub-Saharan Africa have reported the prevalence of LVH among women (mean age range: 43-57 years) to vary from 19.9% to 44.5% [19, 45–47], which is consistent with the current study’s findings (35.8% of South African women; mean age: 41.3 years). Our estimates are, however, higher than previously reported South African data by Maseko et al and Libhaber et al (LVH prevalence of 19.9% and 21.7% respectively, despite similar ages (44 years)) [45, 46]. This increase in LVH prevalence may be, in part, due to increasing risk cardiovascular risk factors from 2013. However, although the populations in both studies were similar to that of the current study (both black South African), the methodology was different. Both studies used LVM indexed to height^2.7^ [45, 46], whilst the current study used LVM indexed to body surface area. However, when indexing LVM to height^2.7^, the current study’s prevalence increased slightly from 35.8% (n=44) to 38.2% (n=47) (data not shown). To date, there remains controversy around the method to index LVM to accurately identify LVH within individuals in the Southern African region. Due to the high prevalence of stunting observed within the South African population [48, 49], and LVM indexed to body surface area was identified as the most appropriate method so as not to under or overestimate LVH calculated by height^2.7^. This, in conjunction with the guidelines from *The American Society of Echocardiography Recommendations for Cardiac Chamber Quantification in Adults* [38], further supports the use of body surface area over height^2.7^.

LVH is a compensatory response to cardiac insult through interactions between pressure and/or volume overload [50]. Increasing age, hypertension, and obesity are the three main cardiovascular risk factors that drive pressure and volume overload within the heart [50, 51]. The current study found that obesity and hypertension were associated with a two-fold and a three-fold respective risk of developing LVH (irrespective of type), and a three-fold risk of developing eccentric LVH. This is in line with results from the Framingham study that suggest a 7% increase for risk for developing LVH per one unit increase of body mass index (BMI) in women [52, 53]. Furthermore, there is a three-fold risk of developing CVD in individuals with BP ≥ 130/90 mmHg compared to individuals with BP < 120/80 mmHg [54]. In South Africa, approximately 68% of women (≥15 years) are overweight or obese [55] and hypertension is currently reported as one of the most common causes of CVD [19, 55]. Current understanding of hemodynamic alterations accompanying obesity and hypertension suggests that they drive an increase in LV filling and stroke volume, which specifically causes LV dilation and eccentric, rather than concentric, LVH [26, 56–59]. However, studies by Woodiwiss et al [60] and Avelar et al [61] found that adiposity was associated with concentric LVH, independently of BP. Our smaller sample size (n=123, of which 11 had concentric LVH) may explain the lack of an association between obesity and concentric LVH in our study. In addition, due to the confounding effects of both hypertension and obesity, their interactions and duration within the same individual left ventricular remodelling and hypertrophy may be different. Therefore, before definitive conclusions can be reached, within this population, further studies investigating interactions of obesity and hypertension are required.

Along with obesity and hypertension, smoking prevalence in South Africa is currently on the rise [55]. There is sound evidence that smoking impacts cardiac structure and function [62, 63], and results from other studies suggest significant increases in LVMI, and thus LVH, in individuals who smoke compared to those that don’t [62, 64, 65]. Similarly, although our sample of women that had concentric LVH was small, our data showed an association between smoking and concentric LVH.

The currently study found a high prevalence of elevated blood pressure (59.4%) in children. Despite this high prevalence it may seem encouraging that only four children had LVH. However, of those four children that had LVH, three had elevated blood pressure and the one child that was normotensive was overweight. Thus, following the same pattern observed in adults, that the rising health risk factors, namely elevated blood pressure and weight gain in children reflect more a pessimistic outlook and are the driving factors behind CVD risk in children that also may persist into adulthood. Further supporting calls for earlier identification of those that are at risk and interventions where necessary, especially within children.

A strength of this study is the indexing of LVM to body surface area. Indexing LVM to body height may result in error in LVH classification due to the prevalence of stunting in this population [48, 49]. A limitation of the present study was the ages within the adult group. As this study recruited from within an existing birth cohort with all index children recruited at birth in 1990, these ‘children’ were all 29 years old adults at the time of the measurements, resulting in a clustering of adults around 29 years with their parents aged 45-65 years. Furthermore, we relied on self-reported data for medical history and health behaviours. Given the potential association between self-reported tobacco use and LVH in adults, analysing biomarkers of tobacco exposure could be a useful next step. As the low frequency of children with LVH prevented further statistical analysis, additional studies with larger sample sizes are needed to assess potential relationships between modifiable health behaviours and LVH.

## Conclusions

In conclusion, the results have highlighted the need for specific call to action polices around obesity, hypertension and tobacco in adult women and target intervention around weight management and elevated BP in children. Due to the high risk of developing LVH due to obesity and hypertension, specific interventions, prior to the development of obesity and hypertension (i.e. overweight and elevated BP individuals), are needed to prevent the development of LVH, and in cases where LVH is evident, regress LVH. As obesity and hypertension have both additive and interactive effects on LVH development, ***both*** weight loss programs and BP lowering medications (such as hydrochlorothiazide [66, 67]) are required to achieve appropriate LVH regression, particular in this population where obesity and hypertension is prevalent.

## Data Availability

The datasets used and/or analysed during the current study are available from the corresponding author on reasonable request

## Abbreviations

ASC: American Society of Echocardiography
BMI: body mass index
BT20+: Birth to Twenty Plus Cohort
BP: blood pressure
CI: confidence interval
CVD: cardiovascular disease
GPAQ: global physical activity questionnaire
IQR: interquartile range
ISH: International Society of Hypertension
IVSd: intraventricular septal diameter
LVDd: left ventricular diastolic diameter
LVPWd: left ventricular posterior wall thickness in diastole
LV: left ventricular
LVH: left ventricular hypertrophy
LVM: left ventricular mass
LVMI: left ventricular mass index
MVPA: moderate-vigorous physical activity
OR: odds ratio
RWT: relative wall thickness
T2DM: type II diabetes mellitus
WHO: world health organisation
WHO-AUDIT: WHO Alcohol Use Disorders
1G: first generation
2G: second generation
3G: third generation

## Declarations

### Ethical approval and consent to participate

Written informed consent was provided by all women and their children / grandchildren prior to their inclusion in the study and ethical approval was obtained from the University of the Witwatersrand’s Research Ethics Committee (Medical) with ethical clearance number **M190263**

### Consent for publication

Not applicable

### Author’s contributions

AKR and LJW, conceptualised and designed the study. AKR was responsible for data management, data analysis and wrote the original draft. AKR, LMS, SHC, SN, WS, SAN, JD, LJW, contributed to interpretation of data and critical review of manuscript. LW was the Principal Investigator of the project. All authors gave final approval of the version to be submitted.

## Acknowledgements

Funding support from the South African Medical Research Council, Wellcome Trust, and DSI-NRF Centre of Excellence in Human Development at the University of the Witwatersrand, Johannesburg, South Africa for the Birth to Twenty Plus Cohort.

